# Association Between Blood Circulating Vitamin D and Colorectal Cancer Risk in Asian Countries: A Systematic Review and Dose-Response Meta-analysis

**DOI:** 10.1101/19000828

**Authors:** Lin Zhang, Huachun Zou, Yang Zhao, Chunlei Hu, Adejare (Jay) Atanda, Xuzhen Qin, Peng Jia, Yu Jiang, Zhihong Qi

## Abstract

**Objectives:** To assess the association between blood circulating Vitamin D levels and colorectal cancer risk in the Asian population.

**Design:** This is a systematic review and dose-response meta-analysis of observational studies that investigated the relationship between blood circulating Vitamin D levels and colorectal cancer risk in the Asian population.

**Data Sources:** Relevant studies were identified through a literature search in MEDLINE, EMBASE, and Web of Science from January 1980 to 31 January 2019. Eligibility criteria: original studies published in peer-reviewed journals investigating the association between blood circulating Vitamin D levels and the risk of colorectal cancer and/or adenoma in Asian countries.

**Data extraction and synthesis:** Two authors independently extracted data and assessed the quality of included studies. Study-specific ORs were pooled using a random-effects model. A dose-response meta-analysis was performed with generalized least squares regression. We applied the Newcastle-Ottawa Scale quality assessment to evaluate the quality of the selected studies.

**Results:** The eight included studies encompassed a total of 2,916 cases and 6,678 controls. The pooled ORs of colorectal cancer for the highest versus lowest categories of blood circulating Vitamin D levels was 0.75 [95% CI, 0.58-0.97] up to 36.5 ng/mL in the Asian population. There was heterogeneity among the studies (*I*^2^=53.9%, *P*_heterogeneity_=0.034). The dose-response meta-analysis indicated a significant linear relationship (*P*_non-linearity_=0.11). An increment of 16 ng/mL in blood circulating Vitamin D level corresponded to an OR of 0.79 [95% CI, 0.64-0.97].

**Conclusions:** The results of this meta□analysis indicate that blood circulating Vitamin D level is associated with decreased risk of colorectal cancer in Asian countries. The dose-response meta-analysis shows that the strength of this association among the Asian population is similar to that among the Western population. Our study suggests that the Asian population should improve nutritional status and maintain a higher level of blood circulating Vitamin D.

**Strengths and limitations of this study:**

- Our study seeks to extend previous work by including a number of new studies and by distinguishing the Asian population explicitly.
- The number of included studies is not sufficient to provide a robust estimate, so the results should be interpreted in the context of the limitations of the available data.
- Heterogeneous definitions of blood circulating Vitamin D categories were used across studies. The variability in definitions could limit comparability between studies.
- Our study included seven case-control studies; the study design implies that the measurement of blood circulating Vitamin D is measured in individuals already diagnosed with colorectal cancer. Results from case-control studies need to be interpreted cautiously because of the potential for reverse causation.
- Time of blood sampling in relation to outcome ascertainment also varied among studies. Such cross-sectional measurements may not accurately reflect an individual’s Vitamin D status across time.

## INTRODUCTION

Colorectal cancer is the third most commonly diagnosed cancer and second in terms of mortality. With over 1.8 million new cases, and 881,000 deaths worldwide in 2018, it accounts for about 1 in 10 cancer cases and deaths.^1^ Some Asian countries where the incidence of colorectal cancer was historically low, such as Japan, Israel, Singapore, China, and the Philippines, have experienced rising incidence rates over the past decades. In 2012, Japan (Miyagi Prefecture Cancer Registry) presented the highest colorectal cancer incidence in the world for men (62 per 100,000 persons) and women (37 per 100,000 persons).^2^ Observational studies have identified several risk factors associated with an increased incidence of colorectal cancer including lifestyle factors (e.g., obesity, physical inactivity, smoking, and heavy alcohol use) and non-modifiable factors (e.g., aging, personal and family history of colorectal cancer or adenoma).^3^ Other observational studies conducted in Western countries suggest blood circulating 25-hydroxyvitamin D (25(OH)D) (Vitamin D) has a protective role in the development of colorectal cancer.^4-8^ Some meta-analyses have consistently reported that there was an inverse association between plasma Vitamin D concentration in the blood and incidence of, and mortality from colorectal cancer.^9-15^

The prevalence of Vitamin D deficiency has increased in recent decades.^16 17^ In a recent population-based study of Asian adults, approximately 75% had suboptimal Vitamin D concentrations.^18^ The Endocrine Society Clinical Practice Guideline defines Vitamin D deficiency as 25(OH)D level <20 ng/mL and insufficiency as 21 to 29 ng/mL.^19^ Feldman et al. ^20^ reported anti-neoplastic actions of Vitamin D, particularly in colorectal cancer.^9^ Touvier et al.^12^ reported that improving Vitamin D levels could be beneficial in reducing colorectal cancer incidence. Data from a cohort of healthy women showed that plasma Vitamin D levels was inversely related to the occurrence and death from colorectal cancer.^21^ In the Nurses’ Health Study, total circulating Vitamin D was associated with a lower risk of colorectal cancer in white women.^22^ A recent international study of 17 cohorts in Western population found that Vitamin D deficiency was associated with increased colorectal cancer risk, and Vitamin D above sufficiency levels was associated with 19% to 27% lower risk.^23^ Compared with Western countries, there was an inconsistent conclusion about the relationship between blood circulating Vitamin D level and colorectal cancer risk in studies of Asian countries,^24-30^ given that lifestyle, ethnic and environmental factors are different between Asian and Western countries.

We hypothesized that the association between blood circulating Vitamin D and colorectal cancer in Asian countries is distinct from Western countries. Thus, this review aimed to summarize epidemiological evidence regarding blood circulating Vitamin D level and colorectal cancer risk in Asian countries. This study underlines the public health importance of attaining and maintaining an optimal vitamin D status in the Asian population and may help to guide clinical and nutritional practice in Asians countries.

## METHODS

We performed the systematic review according to a predetermined protocol and reported in accordance with the Preferred Reporting Items for Systematic Reviews and Meta-Analyses Protocols (PRISMA-P) guidelines.^31^ Two reviewers (L.Z. and Z.Q.) independently undertook the literature search, assessment for eligibility, data extraction, and qualitative assessment. Any inconsistencies between the two reviewers were reviewed by a third reviewer (Y.J.) and resolved by consensus.

### Eligibility criteria

#### Participants

Our study uses the list of sovereign states and dependent territories in Asia by the United Nation (https://unstats.un.org/unsd/methodology/m49/) to draw participants from 48 countries located in five regions (Central Asia, Eastern Asia, Southern Asia, Southeastern Asia, and Western Asia). Asians, and people of Asian origin who live in Western countries were excluded.

#### Exposure

The exposure is blood circulating 25-hydroxyvitamin D (25(OH)D) level which is commonly measured to assess and monitor Vitamin D status in individuals. Most studies only report the total level and do not distinguish D2 and D3 forms of the vitamin. In our meta-analysis, we consider the total level of Vitamin D as the exposure.

#### Comparators (controls)

In order to be eligible for inclusion, studies must compare outcomes in a group of exposed individuals with the highest category of blood circulating Vitamin D level and a group of unexposed individuals with the lowest category of blood circulating Vitamin D level.

#### Outcome

Studies included in the review have a diagnosis of colorectal cancer or colorectal adenoma, clinically confirmed by colonoscopy or pathology.

#### Study design

We target observational studies (case-control, cross-sectional and cohort). English language studies conducted post-1980 were considered eligible. Animal studies were excluded.

Studies were excluded if they: 1) were reviews, editorial, case report, or guideline articles; 2) did not explicitly state the blood circulating Vitamin D level and its association with colorectal cancer risk; 3) allowed controls to have a previous disease history of cancer; 4) focused on Western population or Asian population living in Western countries; or 5) investigated the blood circulating Vitamin D level and its association with survival of colorectal cancer. By consensus among all three reviewers (L.Z., Z.Q. and Y.J.), if data sources were duplicated in more than one study, only the original study was included in the meta-analysis.

### Search strategy

We conducted a literature search using Medline, Embase, and Web of Science, and retrieved all relevant articles that reported the association plasma or serum Vitamin D level and the risk of colorectal neoplasia in Asian countries, published from January 1980 to 31 January 2019. The Medical Subject Heading (MeSH) terms were used in conjunction with the following keywords for our search: (colorectal neoplasm or colon neoplasm or colorectal cancer or colon cancer) AND (25-OH-D or cholecalciferol or calcidiol or calcitriol or 25-hydroxyVitamin D or hydroxycholecalciferols or 25-hydroxyVitamin D3 or 1-alpha-hydroxylase or Vitamin D) AND (Asia* or Afghanistan or Armenia or Azerbaijan or Bahrain or Bangladesh or Bhutan or Brunei or Cambodia or China or Cyprus or Georgia or India or Indonesia or Iran or Iraq or Israel or Japan or Jordan or Kazakhstan or Kuwait or Kyrgyzstan or Laos or Lebanon or Malaysia or Maldives or Mongolia or Myanmar or Burma or Nepal or North Korea or Oman or Pakistan or Palestine or Philippines or Qatar or Russia or Saudi Arabia or Singapore or South Korea or Sri Lanka or Syria or Taiwan or Tajikistan or Thailand or Timor-Leste or Turkey or Turkmenistan or United Arab Emirates or Uzbekistan or Vietnam or Yemen). Full search strings are presented in **Table S1**. References from relevant articles, editorials, conference abstracts, letters, and reviews were thoroughly reviewed to identify additional studies. Full manuscripts of every article with a relevant title and abstract were then reviewed for eligibility.

### Data extraction and qualitative assessment

Two reviewers (L.Z. and Z.H.Q) independently extracted the following study-level characteristics from each eligible study: first author, year of publication, type of study, country where the study was conducted, selection criteria, the numbers of cases and controls (for case-control studies or cross-sectional studies) and the numbers of total participants and incident cases (for cohort studies), population characteristics (sex and age), follow-up period (for cohort studies), sample size, levels of Vitamin D in both case and control group, measures and ranges of Vitamin D, adjusted variables, and risk estimates with corresponding 95% CI for each category. For studies that reported both crude and adjusted estimates of the association between blood circulating Vitamin D and risk of colorectal cancer or adenoma, we used the adjusted estimates for the meta-analysis. For studies that reported several adjusted estimates of association, we used the estimates adjusted for the most variables.

We applied the Newcastle-Ottawa Scale (NOS) quality assessment tool to evaluate the quality of the selected observational studies. This tool was used to measure the key aspects of the methodology in selected studies with regard to design quality and the risk of biased estimates based on three design criteria: 1) selection of study participants; 2) comparability of study groups; and 3) assessment of outcome and exposure with a star system (with a maximum of 9 stars). We judged studies that received a score of 7-9 stars to be at low risk of bias, studies that scored 4-6 stars to be at medium risk, and those that scored 3 or less to be at high risk of bias. A funnel plot was used to assess the publication bias. Any disagreement on the data extraction and quality assessment of the studies were resolved through comprehensive discussion (L.Z., Z.Q, and Y.J.).

### Statistical analysis

Study-specific OR estimates were combined using a random-effects model, that considers within-study and between-study variations. Corresponding 95% CIs were extracted directly from articles where available, with adjusted ORs extracted preferentially over unadjusted ORs. The dose-response analysis was utilized to assess the relationship between blood circulating Vitamin D level and colorectal cancer risk using the generalized least squares (GLS) method to resolve the inconsistency issue of different Vitamin D levels in included studies.^32 33^ For the dose-response meta-analysis of blood circulating Vitamin D levels, we used a method proposed by previous studies to compute the trend from the correlated log OR estimates across categories of Vitamin D levels.^34^ This analytical method collected the distribution of cases and controls, median values of blood circulating Vitamin D levels, and corresponding OR estimates in each category for each study. The assigned value of the lowest category was designated as a reference level. If the study did not provide median values of blood Vitamin D, the midpoint of the upper and lower boundaries in each category was assigned. For the open-ended exposure categories, the length of the open-ended interval was assumed to be the same as that of the adjacent interval. We examined a potential non-linear dose-response relationship between blood circulating Vitamin D level with colorectal cancer risk by modelling Vitamin D levels using random-effect restricted cubic splines with three knots at percentiles 25%, 50%, and 75% of the distribution (spline model). A *P*-value for nonlinearity was calculated by testing the null hypothesis that the regression coefficient of the second spline was equal to zero by Wald-type test of nonlinear hypotheses.^34^ A small *P*-value (<0.05) of the Wald-type test indicates departure from linearity. The non-linear dose-response relationship was confirmed by several representative point values and the risk estimates of a subgroup analysis based on the range of exposure.

The statistical heterogeneity among studies was evaluated using Cochran’s Q test and *I*^*2*^ statistic, with values of 25%, 50%, and 75% representing low, moderate, and high heterogeneity, respectively.^35^ The criterion for identifying heterogeneity was a *P* value less than 0.05 for the Q test. If substantial heterogeneity was detected, we performed univariate meta-regression analyses to explore the proportion of between-study variance explained by study quality, participant characteristics, and study characteristics. We were unable to perform a multivariate meta-regression analysis as only a small number of included studies reported information for all study-level factors. We performed subgroup analyses comparing pooled association estimates and heterogeneity with stratification by, participants sex, outcome type, subregion of Asia, blood sample type, and range of Vitamin D levels (The range is the difference in the midpoint between the highest and lowest categories of blood circulating Vitamin D in each study). An estimation of publication bias was evaluated by the Beggs funnel plot, in which the SE of log (OR) of each study was plotted against its log (OR). An asymmetrical plot suggests possible publication bias. Egger’s linear regression test assessed funnel plot asymmetry, a statistical approach to identify funnel plot asymmetry on the natural logarithm scale of the ORs. All statistical analyses were performed using Stata (version 14.2; StataCorp LP, College Station, Texas). All *P* values were two-sided, and *P* <0.05 was considered as statistically significant.

## RESULTS

### Selection of studies

A detailed PRISMA flow diagram^31^ of literature search and inclusion criteria is shown in **Figure 1**. A total of 611 studies were initially identified with this literature search (250 from Medline, 272 from Embase and 89 from Web of Science), but 114 studies were excluded due to duplication and 465 were excluded after screening the titles and abstracts. Twenty-four other studies were excluded after full-text review (details shown in **Table S2**). Finally, a total of eight studies were identified as eligible for meta-analysis.

**Figure 1.**
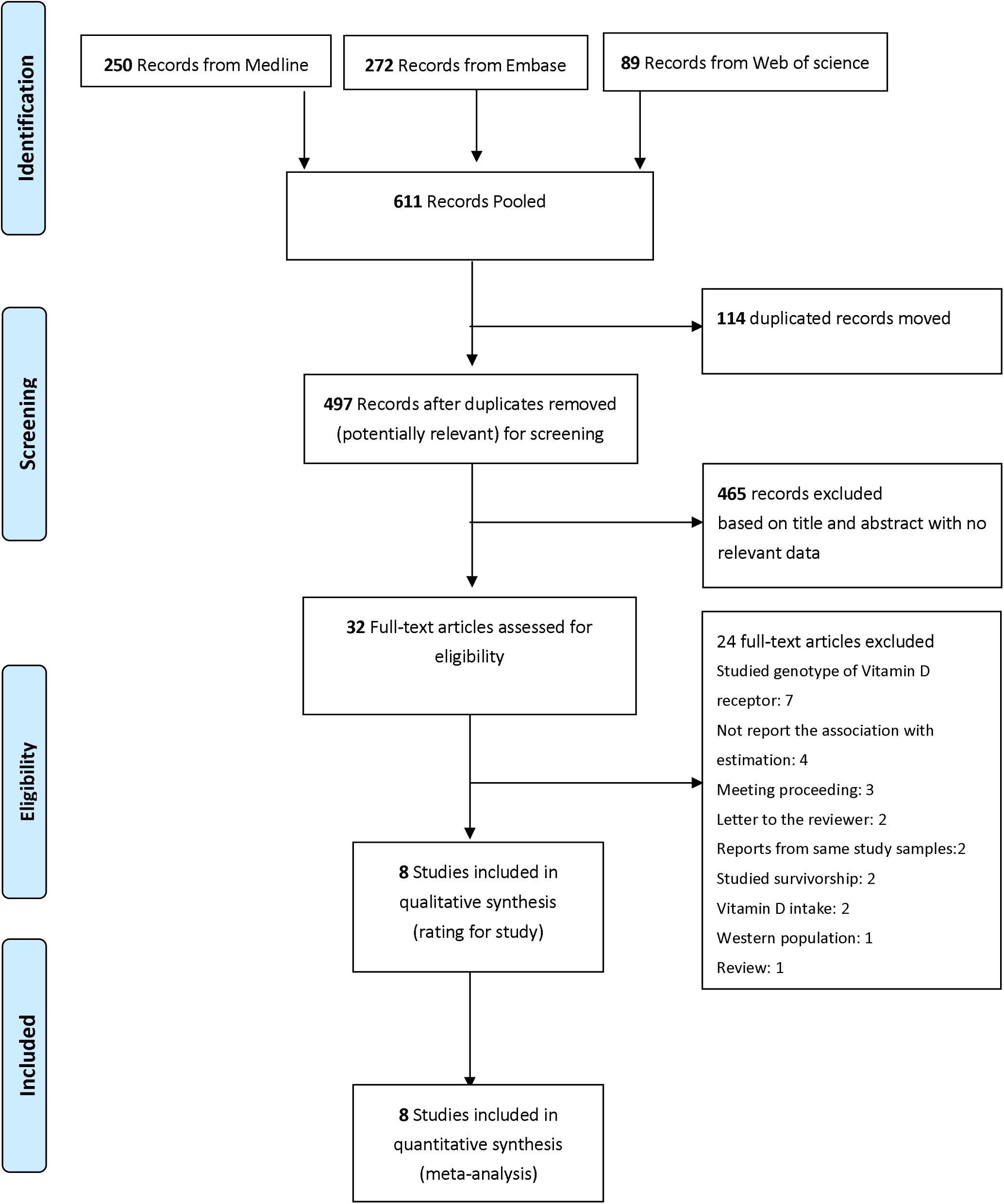
PRISMA flow diagram of study selection for meta-analysis

### Study characteristics

The eight studies included had a total of 2,916 cases and 6,678 controls (**Tables 1 and 2**). These studies were published between 2007 and 2018 - seven from Eastern Asia (four are from Japan,^24 25 27 30^ two from Korea,^26 28^ and one from China;^36^) and one is from Western Asia.^29^ Regarding study design, seven were case-control,^24-29 36^ and one was a nested case-cohort study.^30^ Of the eight studies, four provided the main endpoint of colorectal cancer^24 29 30 36^ and the remaining four provided the main endpoint of colorectal adenoma.^25-28^

**Table 1.**
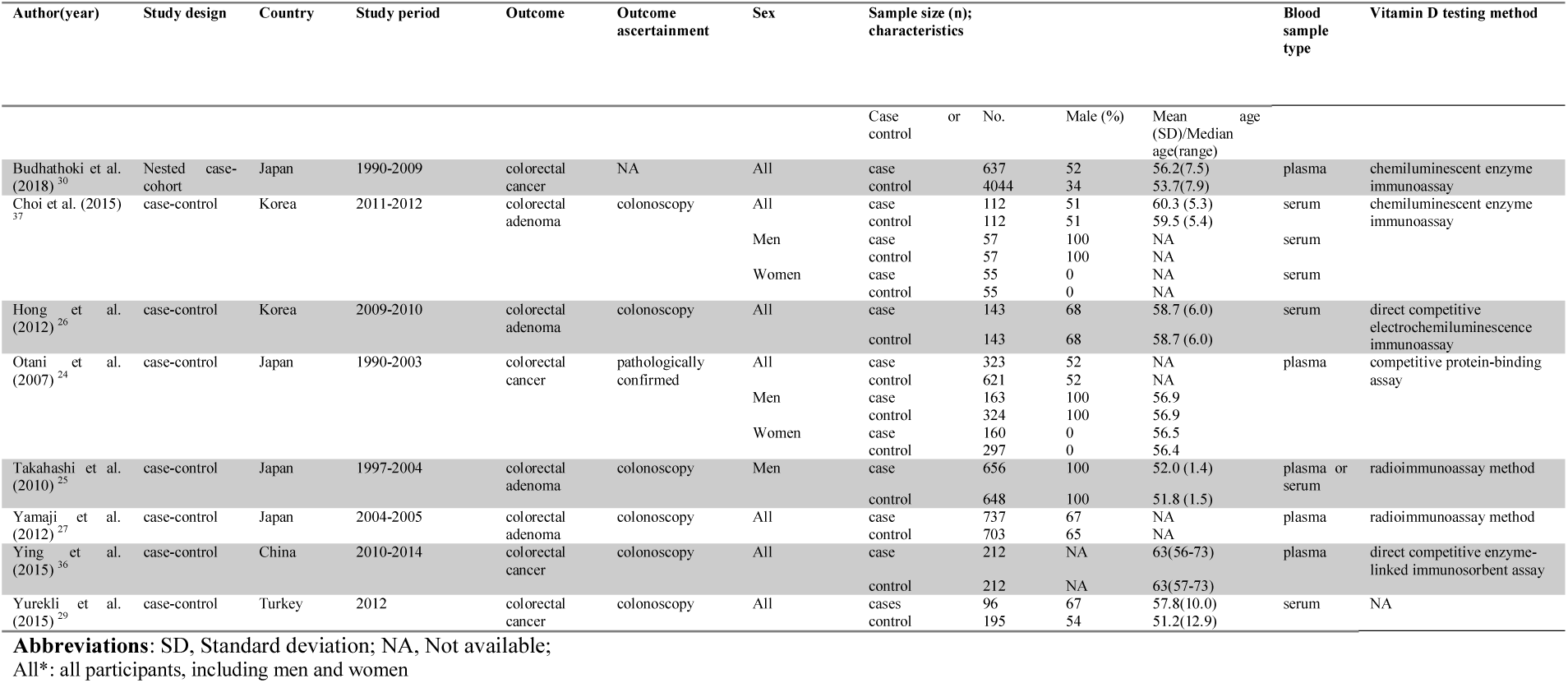
Summary of characteristic for 8 studies included in the meta-analysis

**Table 2.**
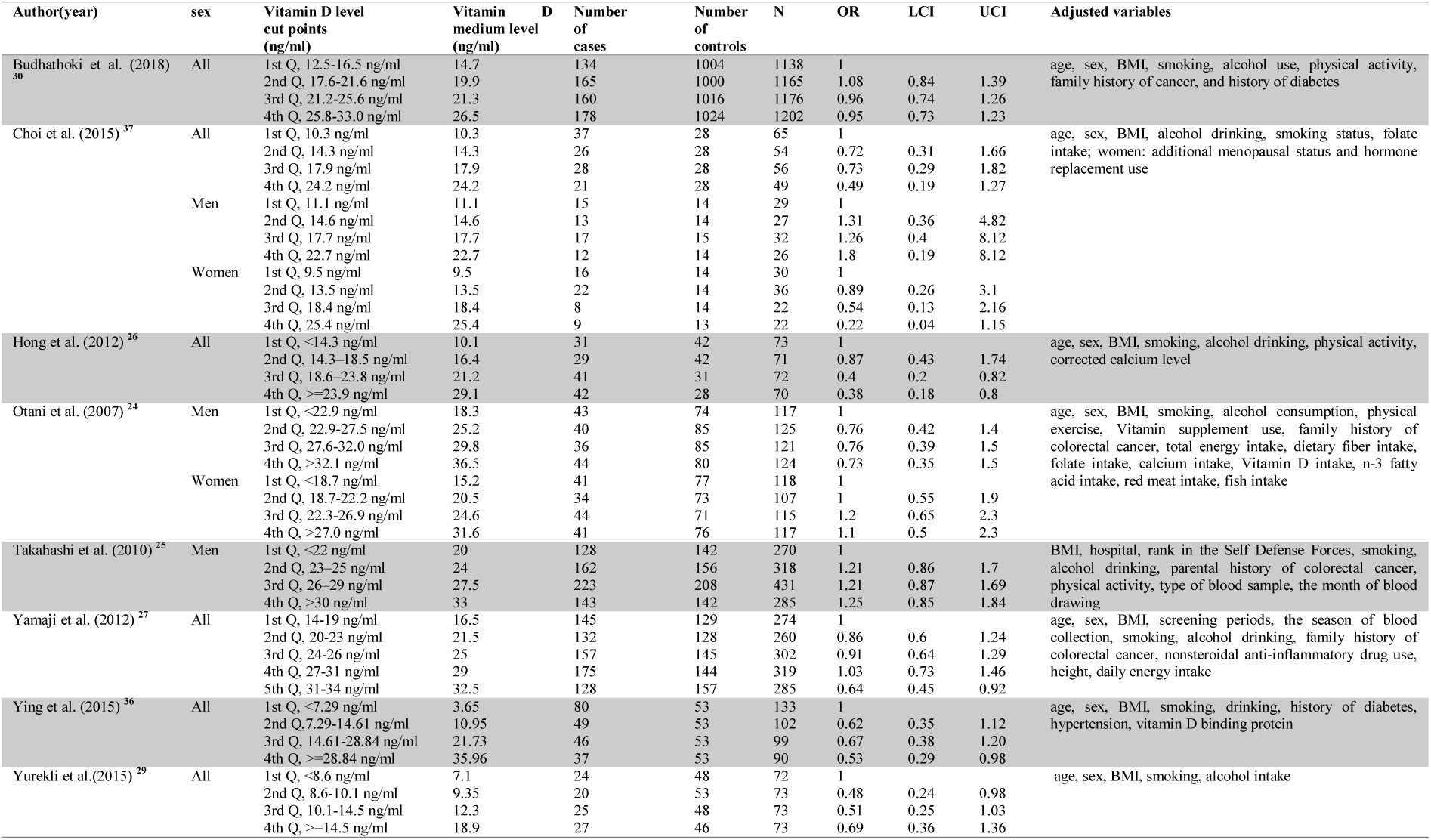
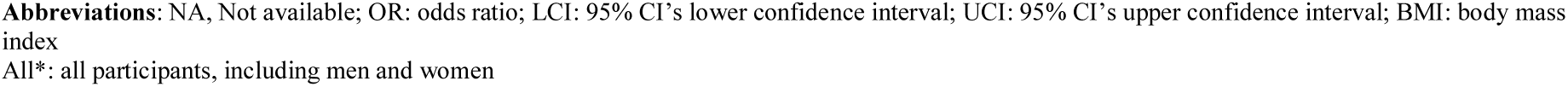
Summary of risk estimate for 8 studies included in the meta-analysis

| Author(year) | sex | Vitamin D level cut points (ng/ml) | Vitamin D medium level (ng/ml) | Number of cases | Number of controls | N | OR | LCI | UCI | Adjusted variables |
| --- | --- | --- | --- | --- | --- | --- | --- | --- | --- | --- |
| Budhathoki et al. (2018)<sup>30</sup> | All | 1st Q, 12.5-16.5 ng/ml | 14.7 | 134 | 1004 | 1138 | 1 |  |  | age, sex, BMI, smoking, alcohol use, physical activity, family history of cancer, and history of diabetes |
|  |  | 2nd Q, 17.6-21.6 ng/ml | 19.9 | 165 | 1000 | 1165 | 1.08 | 0.84 | 1.39 |  |
|  |  | 3rd Q, 21.2-25.6 ng/ml | 21.3 | 160 | 1016 | 1176 | 0.96 | 0.74 | 1.26 |  |
|  |  | 4th Q, 25.8-33.0 ng/ml | 26.5 | 178 | 1024 | 1202 | 0.95 | 0.73 | 1.23 |  |
| Choi et al. (2015)<sup>37</sup> | All | 1st Q, 10.3 ng/ml | 10.3 | 37 | 28 | 65 | 1 |  |  | age, sex, BMI, alcohol drinking, smoking status, folate intake; women: additional menopausal status and hormone replacement use |
|  |  | 2nd Q, 14.3 ng/ml | 14.3 | 26 | 28 | 54 | 0.72 | 0.31 | 1.66 |  |
|  |  | 3rd Q, 17.9 ng/ml | 17.9 | 28 | 28 | 56 | 0.73 | 0.29 | 1.82 |  |
|  |  | 4th Q, 24.2 ng/ml | 24.2 | 21 | 28 | 49 | 0.49 | 0.19 | 1.27 |  |
|  | Men | 1st Q, 11.1 ng/ml | 11.1 | 15 | 14 | 29 | 1 |  |  |  |
|  |  | 2nd Q, 14.6 ng/ml | 14.6 | 13 | 14 | 27 | 1.31 | 0.36 | 4.82 |  |
|  |  | 3rd Q, 17.7 ng/ml | 17.7 | 17 | 15 | 32 | 1.26 | 0.4 | 8.12 |  |
|  |  | 4th Q, 22.7 ng/ml | 22.7 | 12 | 14 | 26 | 1.8 | 0.19 | 8.12 |  |
|  | Women | 1st Q, 9.5 ng/ml | 9.5 | 16 | 14 | 30 | 1 |  |  |  |
|  |  | 2nd Q, 13.5 ng/ml | 13.5 | 22 | 14 | 36 | 0.89 | 0.26 | 3.1 |  |
|  |  | 3rd Q, 18.4 ng/ml | 18.4 | 8 | 14 | 22 | 0.54 | 0.13 | 2.16 |  |
|  |  | 4th Q, 25.4 ng/ml | 25.4 | 9 | 13 | 22 | 0.22 | 0.04 | 1.15 |  |
| Hong et al. (2012)<sup>26</sup> | All | 1st Q, <14.3 ng/ml | 10.1 | 31 | 42 | 73 | 1 |  |  | age, sex, BMI, smoking, alcohol drinking, physical activity, corrected calcium level |
|  |  | 2nd Q, 14.3-18.5 ng/ml | 16.4 | 29 | 42 | 71 | 0.87 | 0.43 | 1.74 |  |
|  |  | 3rd Q, 18.6-23.8 ng/ml | 21.2 | 41 | 31 | 72 | 0.4 | 0.2 | 0.82 |  |
|  |  | 4th Q, >=23.9 ng/ml | 29.1 | 42 | 28 | 70 | 0.38 | 0.18 | 0.8 |  |
| Otani et al. (2007)<sup>24</sup> | Men | 1st Q, <22.9 ng/ml | 18.3 | 43 | 74 | 117 | 1 |  |  | age, sex, BMI, smoking, alcohol consumption, physical exercise, Vitamin supplement use, family history of colorectal cancer, total energy intake, dietary fiber intake, folate intake, calcium intake, Vitamin D intake, n-3 fatty acid intake, red meat intake, fish intake |
|  |  | 2nd Q, 22.9-27.5 ng/ml | 25.2 | 40 | 85 | 125 | 0.76 | 0.42 | 1.4 |  |
|  |  | 3rd Q, 27.6-32.0 ng/ml | 29.8 | 36 | 85 | 121 | 0.76 | 0.39 | 1.5 |  |
|  |  | 4th Q, >32.1 ng/ml | 36.5 | 44 | 80 | 124 | 0.73 | 0.35 | 1.5 |  |
|  | Women | 1st Q, <18.7 ng/ml | 15.2 | 41 | 77 | 118 | 1 |  |  |  |
|  |  | 2nd Q, 18.7-22.2 ng/ml | 20.5 | 34 | 73 | 107 | 1 | 0.55 | 1.9 |  |
|  |  | 3rd Q, 22.3-26.9 ng/ml | 24.6 | 44 | 71 | 115 | 1.2 | 0.65 | 2.3 |  |
|  |  | 4th Q, >27.0 ng/ml | 31.6 | 41 | 76 | 117 | 1.1 | 0.5 | 2.3 |  |
| Takahashi et al. (2010)<sup>25</sup> | Men | 1st Q, <22 ng/ml | 20 | 128 | 142 | 270 | 1 |  |  | BMI, hospital, rank in the Self Defense Forces, smoking, alcohol drinking, parental history of colorectal cancer, physical activity, type of blood sample, the month of blood drawing |
|  |  | 2nd Q, 23-25 ng/ml | 24 | 162 | 156 | 318 | 1.21 | 0.86 | 1.7 |  |
|  |  | 3rd Q, 26-29 ng/ml | 27.5 | 223 | 208 | 431 | 1.21 | 0.87 | 1.69 |  |
|  |  | 4th Q, >30 ng/ml | 33 | 143 | 142 | 285 | 1.25 | 0.85 | 1.84 |  |
| Yamaji et al. (2012)<sup>27</sup> | All | 1st Q, 14-19 ng/ml | 16.5 | 145 | 129 | 274 | 1 |  |  | age, sex, BMI, screening periods, the season of blood collection, smoking, alcohol drinking, family history of colorectal cancer, nonsteroidal anti-inflammatory drug use, height, daily energy intake |
|  |  | 2nd Q, 20-23 ng/ml | 21.5 | 132 | 128 | 260 | 0.86 | 0.6 | 1.24 |  |
|  |  | 3rd Q, 24-26 ng/ml | 25 | 157 | 145 | 302 | 0.91 | 0.64 | 1.29 |  |
|  |  | 4th Q, 27-31 ng/ml | 29 | 175 | 144 | 319 | 1.03 | 0.73 | 1.46 |  |
|  |  | 5th Q, 31-34 ng/ml | 32.5 | 128 | 157 | 285 | 0.64 | 0.45 | 0.92 |  |
| Ying et al. (2015)<sup>36</sup> | All | 1st Q, <7.29 ng/ml | 3.65 | 80 | 53 | 133 | 1 |  |  | age, sex, BMI, smoking, drinking, history of diabetes, hypertension, vitamin D binding protein |
|  |  | 2nd Q, 7.29-14.61 ng/ml | 10.95 | 49 | 53 | 102 | 0.62 | 0.35 | 1.12 |  |
|  |  | 3rd Q, 14.61-28.84 ng/ml | 21.73 | 46 | 53 | 99 | 0.67 | 0.38 | 1.20 |  |
|  |  | 4th Q, >=28.84 ng/ml | 35.96 | 37 | 53 | 90 | 0.53 | 0.29 | 0.98 |  |
| Yurekli et al.(2015)<sup>29</sup> | All | 1st Q, <8.6 ng/ml | 7.1 | 24 | 48 | 72 | 1 |  |  | age, sex, BMI, smoking, alcohol intake |
|  |  | 2nd Q, 8.6-10.1 ng/ml | 9.35 | 20 | 53 | 73 | 0.48 | 0.24 | 0.98 |  |
|  |  | 3rd Q, 10.1-14.5 ng/ml | 12.3 | 25 | 48 | 73 | 0.51 | 0.25 | 1.03 |  |
|  |  | 4th Q, >=14.5 ng/ml | 18.9 | 27 | 46 | 73 | 0.69 | 0.36 | 1.36 |  |

**Abbreviations:** NA, Not available; OR: odds ratio; LCI: 95% CI's lower confidence interval; UCI: 95% CI's upper confidence interval; BMI: body mass index
All\*: all participants, including men and women

### Meta-analysis and dose-response analysis

The multivariable-adjusted ORs for each study and the combination of all eight studies for the highest versus lowest categories of blood circulating Vitamin D levels are shown in **Figure 2**. The mean blood circulating Vitamin D level of the included study population was 20.21 ng/mL, with an SD of 7.92 ng/mL, the minimal concentration of 3.65 ng/mL, and the maximal concentration of 36.5 ng/mL. Results from the studies on blood circulating Vitamin D levels in relation to colorectal cancer risk were inconsistent, with both inverse and positive associations reported. The pooled ORs of colorectal cancer for the highest versus lowest categories of blood circulating Vitamin D level was 0.75 [95% CI, 0.58-0.97], which indicate higher blood circulating Vitamin D level had a significant inverse association with risk of colorectal cancer. There was statistically significant heterogeneity among the studies (*I*^2^=53.9%, *P*=0.034).

**Figure 2.**
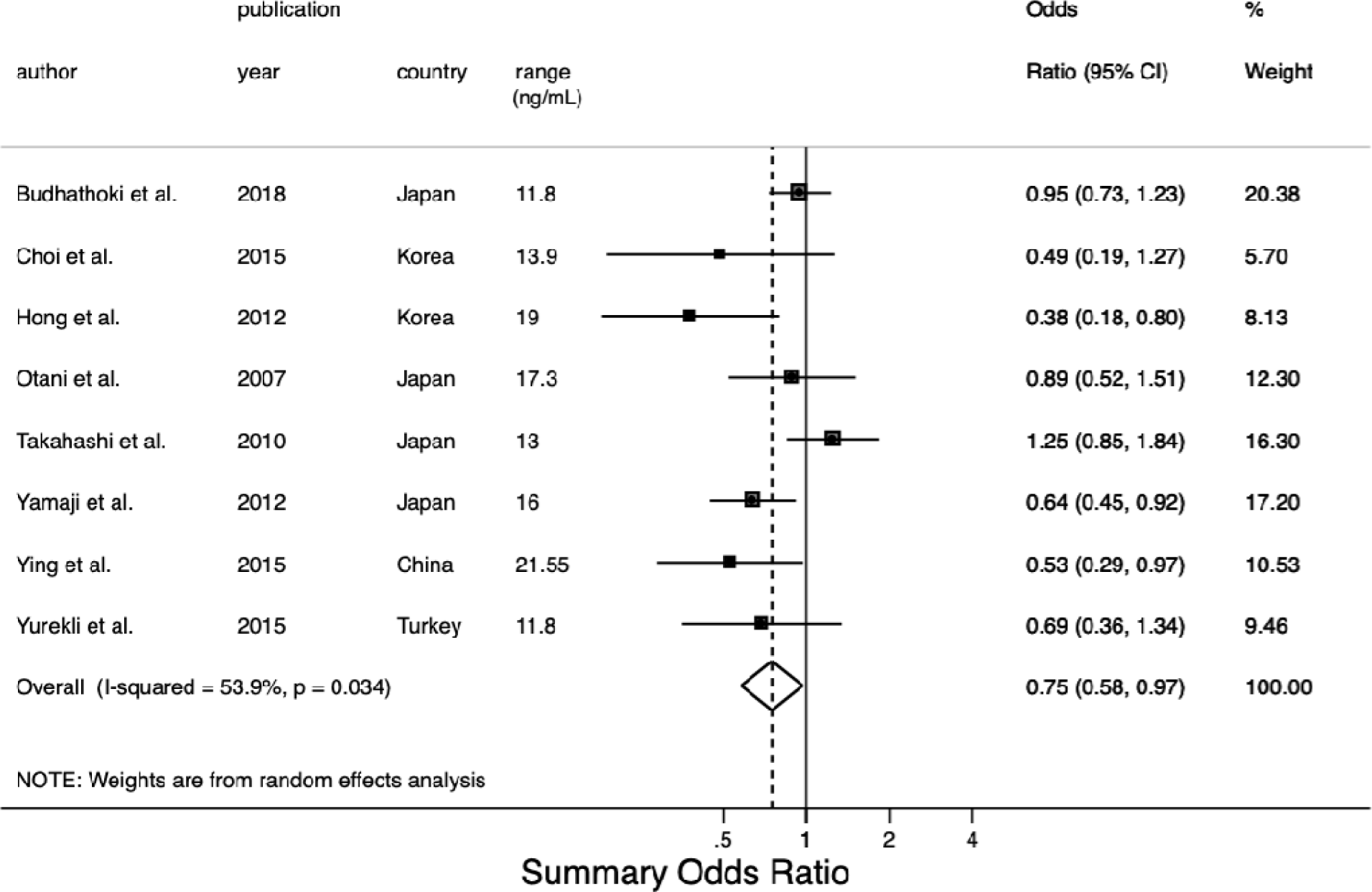
Meta-analysis of association between blood circulating Vitamin D and the risk of colorectal cancer. The adjusted odds ratios were included of colorectal cancer for the highest versus lowest categories of blood Vitamin D level from each study. The size of each square is proportional to the weight of the study. The range is the difference in the midpoint between the highest and lowest categories of blood Vitamin D. F, female; M, male;

We evaluated the non-linear dose-response relationship between blood circulating Vitamin D levels and colorectal cancer risk. A 16 ng/mL increment (about 2 SDs=15.84 ng/mL) in blood circulating Vitamin D levels conferred an OR of 0.79 [95% CI, 0.64-0.97], which meant the risk of colorectal cancer decreased by 21% for every 16 ng/mL increment in blood Vitamin D, and the reduction was statistically significant. A moderate heterogeneity existed (*I*^2^=53.9%, *P*_heterogeneity_=0.034) in the overall analysis of blood circulating Vitamin D levels, without a significant non-linear dose-response relationship (*P*_non-linearity_=0.11), suggesting that the non-linear dose-response relationship does not depart from linearity. Similar trends were observed with linear and spline models (**Figure 3**).

**Figure 3.**
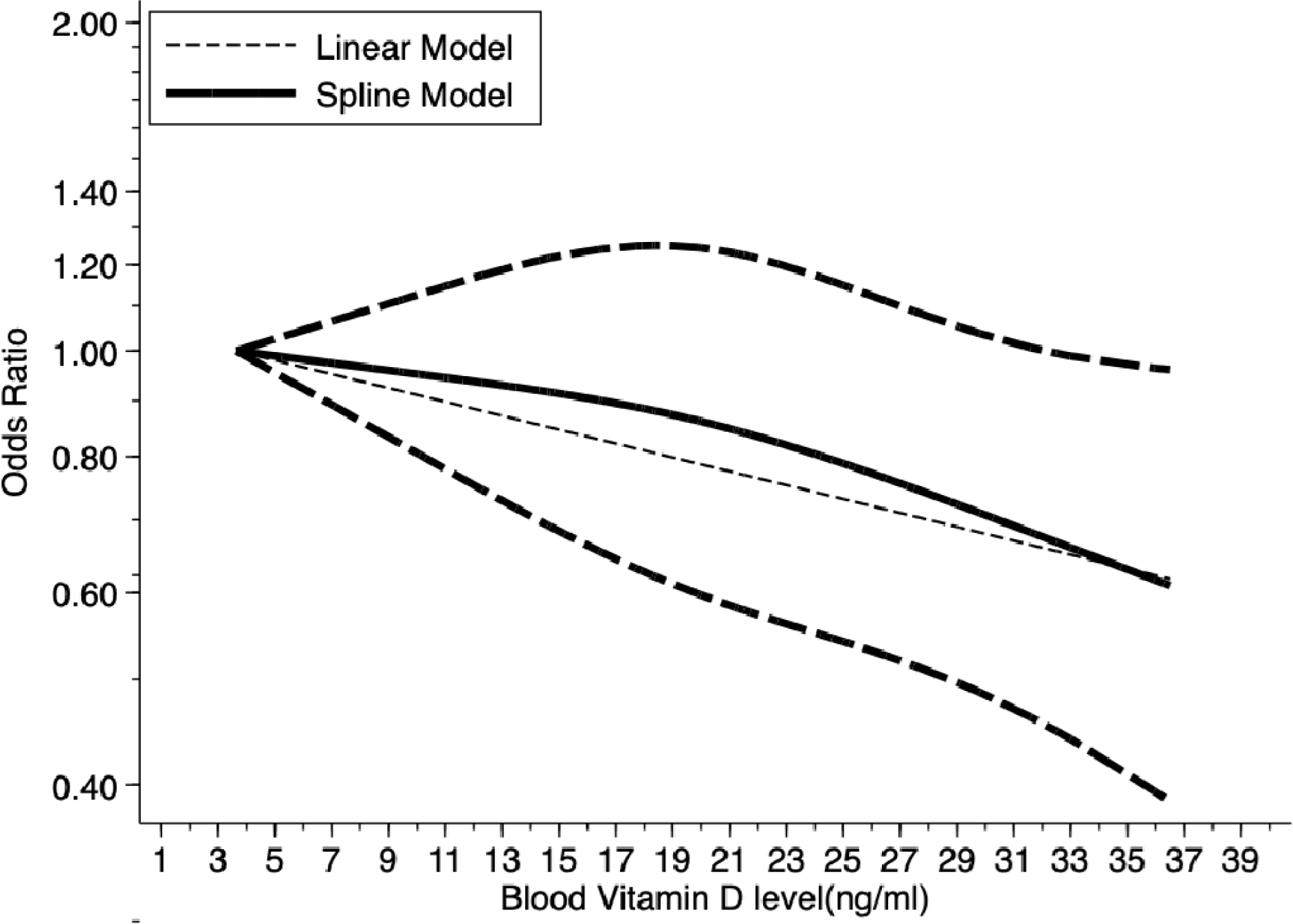
Dose-response relationship between blood Vitamin D levels and the ORs of colorectal cancer. Adjusted Odds Ratio and 95% CIs (dashed lines) are reported. Blood Vitamin D levels were modelled with a linear and spline model in a random-effects meta-regression model. The median value of the lowest reference interval (3.65 ng/mL) was used to estimate all Odds ratios. The vertical axis is on a log scale. Lines with long dashes represent the pointwise 95% confidence intervals for the fitted spline model with a nonlinear trend (solid line, *P*_non-linearity_=0.11). Lines with short dashes represent the linear trend.

### Subgroup analysis

When we stratified the analysis according to blood sample type, the pooled ORs of serum sample and plasma for the highest versus lowest categories of blood circulating Vitamin D levels were 0.52 [95% CI, 0.34-0.80] and 0.77 [95% CI, 0.59-1.00] respectively. The results showed that there was a substantial risk reduction (48%) for blood serum Vitamin D levels associated with the risk of colorectal cancer. There was no evidence of significant statistical heterogeneity among studies (*I*^2^=22.67%, *P*=0.21). We then performed a subgroup analysis of blood circulating Vitamin D range in each study. The pooled ORs was 0.93 [95% CI, 0.70-1.25] for studies with range ≤15 ng/mL and 0.62 [95% CI, 0.47-0.83] for studies with range >15 ng/mL. There was no evidence of statistical heterogeneity among studies (*I*^2^=28.48%, *P*=0.08).When stratified by outcome, the pooled ORs were 0.67 [95% CI, 0.40-1.14] for studies when the outcome was colorectal adenoma and 0.83 [95% CI, 0.66-1.06] when the outcome was colorectal cancer respectively with no significantly statistical heterogeneity among studies (*I*^2^=59.48%, *P*=0.73). When we stratified the studies by sex, the pooled ORs were 0.59 [95% CI, 0.13-2.74] for studies with estimates for women, and 1.13 [95% CI, 0.81-1.58] for men respectively with no significantly statistical heterogeneity among studies (*I*^2^=45.89%, *P*=0.70). We also stratified according to the geographical region of the population. The pooled ORs was 0.75 [95% CI, 0.57-1.00] for Eastern Asia and 0.69 [95% CI, 0.36-1.34] for Western Asia. There was no significantly statistical heterogeneity among studies (*I*^2^=59.79%, *P*=0.87). The subgroup meta-analyses are shown in **Figure S1** and summarized in **Table 3**.

**Table 3.** Summary of subgroup analysis for the associations of blood circulating Vitamin D and the risk colorectal cancer and adenoma by outcome, sex, the subregion of Asia, and blood sample type.

| Subgroup analysis | Number of estimates | Random effect The summary OR (95% CI) | Ratio of ORs (95% CI) | $I^2$ (%) | $P$ value |
| --- | --- | --- | --- | --- | --- |
| **Outcome** |  |  |  |  |  |
| Colorectal adenoma | 4 | 0.67 (0.40-1.14) |  |  |  |
| Colorectal cancer | 4 | 0.83 (0.66-1.06) | 1.11(0.54-2.28) | 59.48 | 0.73 |
| **Sex** |  |  |  |  |  |
| Women | 2 | 0.59 (0.13-2.74) |  |  |  |
| Men | 3 | 1.13(0.81-1.58) | 1.46 (0.03-63.5) | 45.89 | 0.70 |
| **Subregion** |  |  |  |  |  |
| Eastern Asia | 7 | 0.75 (0.57-1.00) |  |  |  |
| Western Asia | 1 | 0.69 (0.36-1.34) | 0.92 (0.28-2.99) | 59.79 | 0.87 |
| **Blood sample type** |  |  |  |  |  |
| Serum | 3 | 0.52 (0.34-0.80) |  |  |  |
| Plasma | 4 | 0.77 (0.59-1.00) | 1.49 (0.73-3.03) | 22.67 | 0.21 |
| **Range** |  |  |  |  |  |
| ≤15 ng/mL | 4 | 0.93 (0.70-1.25) |  |  |  |
| >15 ng/mL | 4 | 0.62 (0.47-0.83) | 0.65(0.39-1.07) | 28.48 | 0.08 |
| Abbreviations: OR, Odds ratio; $I^2$, Percent residual variation due to heterogeneity; CI, Confidence interval. | | | | | |
| *$I^2$ and P value related to subgroup differences. | | | | | |

### Qualitative assessment and publication bias

The NOS tool was used to conduct a qualitative assessment of the selected studies to review the quality of the studies and detect possible bias. Of the eight studies, five were at low risk of bias (8-9) stars.^24-27 30^ Three studies were at medium risk (5-7 stars) mainly due to bias from representativeness of cases or controls, control definition, and response rate.^29 36 37^( shown in **Table S3**.) The funnel plot and Eggers statistical test indicated no evidence of publication bias in the studies included in the meta-analysis (*P*=0.338) (**Figure S2**.)

## DISCUSSION

Colorectal cancer is one of the cancers with high morbidity and mortality in the world. The one-carbon metabolism pathway requires adequate Vitamin D, and this raises the possibility that Vitamin D may have an essential role in the risk of colorectal cancer. Many epidemiological studies from Europe and the United States believe that increasing the concentration of circulating Vitamin D can reduce the morbidity and mortality of colorectal cancer.^38^ However, the association between blood circulating Vitamin D levels and the risk of colorectal cancer in the Asian population is still under debate due to a lack of sufficient evidence. This systematic review highlights the inconsistencies among studies addressing the role of blood circulating Vitamin D and colorectal cancer risk in the Asian population. Our systematic review identified eight studies consisting of 2916 cases and 6678 controls that addressed the relationship between blood circulating Vitamin D levels and colorectal cancer risk. Our meta-analysis found 25% reduced risk of colorectal cancer for the highest versus lowest categories of blood circulating Vitamin D levels (OR=0.75, 0.58-0.97) up to 36.5 ng/mL, that indicated higher blood circulating Vitamin D level has a significant inverse association with risk of colorectal cancer in Asian population. Our meta-analysis results showed that the negative correlation between Vitamin D and the risk of colorectal cancer is similar to that of European and American population studies^11 12 21 22 39-42^ and consistent with the result of a meta-analysis by Ekmekcioglu C et al.^13^, that found a pooled relative risk (RR) of 0.62 (0.56–0.70) for colorectal cancer when comparing individuals with the highest category of 25(OH)D with those in the lowest.

Our results found a 16 ng/mL increment in blood circulating Vitamin D levels with an OR of 0.79 [95% CI, 0.64-0.97], which meant the risk of colorectal cancer decreased by 21% for every 16 ng/mL increment in blood Vitamin D, and the reduction was statistically significant. Our study also suggested a linear dose-response relationship (*P*_non-linearity_= 0.11), that was consistent with several studies conducted in Western populations revealing a similar protective dose-response association of the blood circulating Vitamin D and colorectal cancer risk. For example, a meta-analysis reported a 26% lower risk of colorectal cancer per 10 ng/mL increment in blood circulating Vitamin D levels.^11^ Most experts define Vitamin D deficiency as a Vitamin D level of less than 20 ng/mL.^43 44^ Vitamin D concentration of 21 to 29 ng/mL can be considered to indicate a relative insufficiency of Vitamin D, and a level of 30 ng/mL or higher can be considered to indicate sufficient Vitamin D.^45^ With the use of such definitions, it has been estimated that many people have Vitamin D deficiency or insufficiency.^43 44^ Previous research has implied an association between Vitamin D deficiency and an increased incidence of bone fractures.^46^ There is also data showing Vitamin D deficiency to be associated with cancer,^47-49^ diabetes,^50^ cognitive impairment,^51^ and all-cause mortality.^52^ Among colorectal cancer patients, the prevalence of Vitamin D deficiency was much higher (nearly 90%) than among patients with other chronic diseases.^53^ Humans obtain Vitamin D from exposure to sunlight, from natural diets, fortified diets, supplementation etc.^46^ Dose-response between Vitamin D concentrations and colorectal cancer risk may be different between Asian and Western populations due to ethnic, anthropometric, dietary, and environmental factors.^12^ Lifestyle and diet can promote the development of early-onset colon lesions by regulating growth factors that interact with inflammatory pathways.^41^ An association between Vitamin D status and reduced risk of colorectal cancer has been found in ethnically diverse populations.^5^ Vitamin D interacts with calcium to enhance the reduction of colon cancer risk.^54-56^ Studies have shown that Vitamin D and calcium may interact and that both are needed in reducing cancer risk.^57^ However, even after adjusting for calcium intake in some studies,^6 58^ Vitamin D was associated with a lower risk. The independent effects of Vitamin D are supported, but the combined effects of Vitamin D and calcium may be greater than the sum of their independent effects.^59^ Vitamin A has an antagonistic effect on Vitamin D ^58^ and taking both at the same time can lead to decreased calcium absorption. Still, Vitamin A is often combined with Vitamin D in supplements.

Vitamin D supplementation significantly reduced total cancer mortality but did not reduce total cancer incidence. ^60^ Patients with low-risk prostate cancer under active surveillance may benefit from vitamin D3 supplementation at 4000 IU/d.^61^ Among patients with metastatic colorectal cancer, the addition of high-dose Vitamin D3, vs standard-dose Vitamin D3, to standard chemotherapy resulted in a difference in median progression-free survival that was not statistically significant, but with a significantly improved supportive effect.^62^ Epidemiological evidence links the incidence of colorectal cancer to lifestyle, smoking, physical activity, alcohol consumption, and sleep.^63^ It has also been linked to reduced fruit and vegetable consumption and increased consumption of red meat. Dairy products, fish and other foods and cooking methods also play an essential role.^64^ In addition, some drugs such as non-steroidal anti-inflammatory drugs and cyclooxygenase inhibitors are also involved.^65^ Women on estrogen therapy, for example, did not reduce their risk of colorectal cancer by taking Vitamin D and calcium supplements.^66^ Increased dietary fiber intake reduces the risk of colorectal cancer and obesity; and low physical activity may reduce plasma 25(OH)D concentration, thereby increasing the risk of colorectal cancer.^42^ For every 1kg/m^2^ increase in BMI of colorectal cancer patients, serum Vitamin D level decreased significantly (0.46 ng/mL).^67^ Most variation in Vitamin D levels usually comes from exposure to the sun, which is an essential source of Vitamin D for people who get more from fish, even in Japan.^68^

This meta-analytic comparison revealed a statistically significant beneficial effect of blood circulating Vitamin D for colorectal cancer. However, the diversity of the studies and the presence of moderate heterogeneity in the overall analysis of blood circulating Vitamin D levels (*I*^2^=53.9%, *P*_heterogeneity_=0.034) may preclude making meaningful conclusions from the pooled estimate because it may not reflect the true underlying effect. The subgroup analysis did not explain much of the heterogeneity.

Potential reasons for the heterogeneity in the strength of the association may include the following. Firstly, food consumption and vitamin supplements varied according to the specific dietary habits and lifestyle in each Asian subregion. A systemic review studied correlations between various diet types, food or nutrients and colorectal cancer risk among Asians, and suggested that red meats, processed meats, preserved foods, saturated/animal fats, cholesterol, high sugar foods, spicy foods, tubers or refined carbohydrates have a positive association with colorectal cancer risk.^69^ Besides diet, other personal and lifestyle factors (e.g., exposure to sunlight, obesity, smoking and drinking habit) may alter the strength of the association and contribute to the heterogeneity of the association in Asian countries. The numbers of studies from the different Asian subregions included in our meta-analysis was imbalanced, so we were cautious in interpreting results. For example, our meta-analysis included seven studies from Eastern Asia, only one from Western Asia, and none from Central, Southern or South-eastern Asia. Our study revealed no evidence of publication bias, and most of the studies included in our meta-analysis verified the diagnosis of colorectal cancer for cases. Histologic confirmation of cancer diagnoses for cases was an optimal validation for the case-control design in our meta-analysis.

Possible confounders for the association between colorectal cancer and blood circulating Vitamin D levels include sex, age, family history of colorectal cancer, smoking, alcohol drinking, body mass index, and diabetes. Most studies in our meta-analysis provided risk estimates that were adjusted for age,^24 26-30 36^ sex,^24 26 27 29 30 36^ body mass index,^24-30 36^ smoking,^24-30 36^ drinking,^24-26 29 36^ physical activity,^30^ alcohol consumption,^24-30^ and family history of colorectal cancer.^24 25 29^ Fewer were adjusted for folate,^24 37^ energy intake,^24 27^ hypertension,^36^ vitamin D binding protein,^36^ blood collection,^24 27^ or for vitamin supplement use.^24^ For these studies, the observed reduced risk of colorectal cancer associated with Vitamin D levels is likely confounded by one or more of these factors.

In the overall analysis for both adenoma and carcinoma that is part of our subgroup analysis, we report a statistically significant association; yet, in the stratified analysis by colorectal adenoma or colorectal cancer separately, the association was not statistically significant. The results, however, show that associations were in the same direction (ORs<1 indicating an inverse relationship). Uncontrolled confounders (e.g. dietary sources of Vitamin D, consumption of fish/fibres containing 25(OH)D, exposure to the sun, folate/calcium intake, Vitamin D supplement use etc.) in the original studies that are part of our meta-analysis may be responsible for these differences. Of note, only one of the 8 studies in our meta-analysis adjusted for these confounders.^24^ The presence of negative confounders in original colorectal cancer studies (OR’s are closer to 1), and positive confounders in original colorectal adenoma studies (OR’s are further from 1) as well as some effect modification may also be responsible for the statistical significance difference noted. We used a random effects model in our analysis to reduce this effect. Further investigation of the subgroup analysis show that the weight of the studies could also be contributory. Two studies^25 30^ contribute a large weight in the subgroup meta-analysis, but a smaller weight in the overall analysis.

Our analysis had the following limitations. First, the number of included studies is not sufficient to provide a robust estimate of the association of blood circulating Vitamin D levels and colorectal cancer risk, so the analysis and results should be interpreted in the context of the limitations of the available data. Second, heterogeneous definitions of blood circulating Vitamin D categories were used across studies. The variability in definitions could limit comparability between studies. Third, our study included seven case-control studies; the study design impling that the measurement of blood circulating Vitamin D is in individuals already diagnosed with colorectal cancer. The results from this study design need to be interpreted cautiously because of the potential for reverse causation. The time of blood sampling in relation to outcome ascertainment also varied among studies. Such cross-sectional measurements may not accurately reflect an individual’s Vitamin D status across time. Fourth, some studies included in our meta-analysis did not adjust for potentially relevant confounders which may have led to residual confounding and may explain some of the observed heterogeneity. Fifth, the difference in the method used for measuring blood circulating Vitamin D levels may also be a source of heterogeneity between the included studies. Sixth, in the dose-response analysis, the literature selected listed the median Vitamin D value, instead of the original Vitamin D value, which could also lead to inaccurate results. Seventh, although we assessed the quality of the observational studies in this meta-analysis with the NOS quality assessment tool, Bae ^70^ suggested that it is more reasonable to control for quality level by performing subgroup analysis according to study design rather than by using the NOS tool. In this context, however, we did not perform subgroup analysis according to study design since the included 8 studies were case-control and nested case-cohort studies. Finally, a recent meta-analysis investigated the association between blood circulating Vitamin D levels and survival of colorectal cancer and found that the pooled hazard ratios (95% confidence intervals) were 0.68 [0.55–0.85] and 0.67 [0.57–0.78] respectively for overall and colorectal cancer-specific survival comparing highest versus lowest categories of blood Vitamin D.^15^ Our study did not explore the association of blood circulating Vitamin D levels and colorectal cancer mortality however, and this association in Asian countries is one we would encourage future studies to examine.

## CONCLUSION

Blood circulating Vitamin D levels is inversely associated with colorectal cancer prevalence in the Asian population. Our findings on the inverse association between blood circulating Vitamin D and the risk of colorectal cancer in the Asian population suggest the need for Asians to improve their nutritional status and maintain higher blood circulating Vitamin D levels. This meta-analysis provides valuable information for future research on the association between blood circulating Vitamin D and colorectal cancer risk in the Asian population. A multinational, population-based study in Asian countries may resolve the issue of heterogeneity and generate detailed information on blood circulating Vitamin D levels and the risk of colorectal cancer. Further studies may also focus on evaluating the association of Vitamin D levels with colorectal cancer mortality.^60^

## Data Availability

All data relevant to the study are included in the article or uploaded as supplementary information.

## Authors Contributions

LZ: Study concept and design; acquisition of data; statistical analysis; interpretation of data; drafting and critical review of the manuscript for important intellectual content; approval of the final version of the manuscript.

HZ: Interpretation of data; drafting and critical review of the manuscript for important intellectual content; approval of the final version of the manuscript.

YZ: Interpretation of data; drafting and critical review of the manuscript for important intellectual content; approval of the final version of the manuscript.

CH: Drafting and critical review of the manuscript for important intellectual content; approval of the final version of the manuscript.

AA: Drafting and critical review of the manuscript for important intellectual content; approval of the final version of the manuscript.

XQ: Interpretation of data; drafting and critical review of the manuscript for important intellectual content; approval of the final version of the manuscript.

PJ: Interpretation of data; drafting and critical review of the manuscript for important intellectual content; approval of the final version of the manuscript.

YJ: Study concept and design; acquisition of data interpretation of data; drafting and critical review of the manuscript for important intellectual content; approval of the final version of the manuscript.

ZQ: Study concept and design; acquisition of data; interpretation of data; drafting and critical review of the manuscript for important intellectual content; approval of the final version of the manuscript.

